# Repeatability of gait of children with spastic cerebral palsy in different walking conditions

**DOI:** 10.1101/2024.04.10.24304953

**Authors:** Laure Everaert, Tijl Dewit, Catherine Huenaerts, Anja Van Campenhout, Luc Labey, Kaat Desloovere

**Affiliations:** KU Leuven Department of Rehabilitation Sciences, Leuven, Belgium; Clinical Motion Analysis Laboratory, University Hospitals Leuven, Leuven, Belgium; KU Leuven, Locomotor and Neurological Disorders, Department of Development and Regeneration, Leuven, Belgium; University Hospitals Leuven, Department of Orthopedics, Leuven, Belgium; KU Leuven, Department of Mechanical Engineering, Geel, Belgium

## Abstract

**Objectives:** Three-dimensional gait analysis is the ‘gold standard’ for measurement and description of gait. Gait variability can arise from intrinsic and extrinsic factors and may vary between walking conditions. This study aimed to define the inter-trial and inter-session repeatability in gait analysis data of children with cerebral palsy (CP) who were walking in four conditions, namely barefoot or with ankle-foot orthosis (AFO), and overground or treadmill.

**Design:** Test-retest repeatability study.

**Setting:** Rehabilitation facility with a human motion analysis laboratory.

**Participants:** Ten ambulatory children with spastic CP (age=5-15years). In addition, two control datasets (N=56; N=18) of typically developing children were used as a reference.

**Interventions:** Not applicable.

**Main Outcome Measures:** Inter-trial and inter-session variability was measured using intra-class correlations (ICC’s) with accompanying confidence intervals, standard error of measurement (SEM), as well as the SEM expressed as percentage (%SEM) of the total joint-range-of-motion of typically developed children.

**Results:** Overall we found good to excellent ICC-values and favourable SEM-values for the inter-session Gait Profile Score (ICC=0.85-0.98, SEM=0.45°-0.91°) and Gait Variable Scores (ICC=0.85-0.99, SEM=0.22-1.11°) for the lower-limb joints. The %SEM was the highest for the ankle joint (%SEM=0.8%–3.0%). For the continuous waveform data, only in the ankle joint, differences were observed. Namely, smaller SEM-values for the AFO-condition (mean inter-trial=0.14°; mean inter-session=1.121°) in comparison to the barefoot-condition (mean inter-trial= 0.55°; mean inter-session=2.22°). For all the kinetic parameters, the treadmill conditions showed smaller SEM-values in comparison to the overground condition.

**Conclusions:** All conditions proved to be repeatable, showing good to excellent ICC-values. The ankle kinematics were more repeatable when the participants were walking with their AFOs in comparison to barefoot walking. Taking the total joint-range-of-motion into account, the knee joint showed the most repeatable motion, while ankle motions showed the lowest repeatability. For kinetics, treadmill conditions showed better repeatability than the overground conditions.

## Introduction

With a prevalence of 1.48 per 1000 live births, cerebral palsy (CP) is the most common cause of permanent motor disabilities in childhood^1^. Three-dimensional gait analysis (3DGA) is a common assessment within the clinical follow-up of children with cerebral palsy (CP) and is the ‘gold standard’ for measurement and description of gait^2^. The 3DGA provides the clinicians with an extended and complex dataset. Therefore, specific gait indices have been developed, which describe the quality of the gait pattern in a single score, such as the gait profile score (GPS) and the gait variable score (GVS)^3^. However, such data reduction techniques ignore many details of the gait pathology. Hence, the use of complete gait cycle waveforms for interpretation of pathological gait gained popularity, which has been facilitated by the uprise of statistical parametric mapping as an established analysis technique in gait-related research^4^. Individual variability can be observed in these gait waveforms and should be considered when interpreting the results. Variations in 3DGA-data can arise naturally (intrinsic) or from experimental errors (extrinsic)^5^. Intrinsic variability cannot be avoided, but provides a baseline estimation of variation in a specific walking condition (such as barefoot walking or walking with orthoses and overground walking and walking on a treadmill), while extrinsic variability can be targeted by quality improvement measures^5^. The variation observed across multiple sessions of the same individual is crucial in the context of clinical gait analysis, and can be investigated by conducting repeated observations over a period of time. Clinical gait analysis forms the basis for adjusting the patient’s clinical care (i.e., botulinum toxin injections, surgery, etc.). If experimental errors mask relevant gait deviations, valuable information will go unnoticed.

The current study was triggered by the lack of knowledge on the repeatability of gait in children with CP. First, there is an overall lack of studies that focused on the repeatability of gait waveforms in disabled children. Indeed, the majority of previous repeatability studies have been performed on adults and/or on typically developing children^6^. Secondly, previous studies were always carried out in the barefoot condition^6,7^. However, in practice, ankle-foot orthoses (AFOs) are often prescribed for improving gait in this population, especially for children who are classified as levels I-III on the gross motor function classification system (GMFCS)^8^. Understanding the repeatability of gait for children walking in AFO-conditions is crucial, given that 3DGA is frequently employed to assess the impact of AFOs on gait. Thirdly, the majority of earlier studies conducted on children focused exclusively on characterizing gait in an overground setting, as traditional 3DGA is typically carried out on an overground walkway^7,9^. However, treadmill 3DGA has gained popularity, since the treadmill condition allows to evaluate gait over a longer period, it reduces the needed square meters for a gait lab and creates opportunity of using virtual reality feedback. Consequently, there is a need for repeatability studies investigating this latter condition.

The current study was set-up to fill in these gaps in the literature concerning repeatability of gait in children with CP. The aim of this study was to determine the repeatability of gait waveforms as well as gait deviation indices in a cohort of children with CP who were walking in four different conditions (barefoot or with AFOs, both on the treadmill or overground). To achieve the overall aim, the following three specific sub-questions were addressed. (a) What is the repeatability of gait in overground barefoot condition in children with CP? (b) What is the repeatability of barefoot walking compared to walking with AFOs? (c) What is the repeatability of barefoot walking in an overground walkway compared to a treadmill condition?

## Methods

### Participants

Prior to the study, the sample size was determined based on the approximations of Bujang et al. (2017)^10^ . For two observations of the subject with an intra-class correlation of 0.7^11^, a power of 80% and alpha of 0.05, a sample size of ten participants was considered sufficient to define relative reliability.

Ten children with spastic CP (7 ⍰; 9.9y ± 3.5y; GMFCS-level I (N=4), level II (N=3) and level III (N=3); uni-(N=4) and bilateral (N=6) CP) were included in this study. Exclusion criteria were (a) severe contractures or spasticity making it impossible to wear a conventional AFO and/or (b) cognitive or visual impairments and/or (c) bone and/or soft-tissue surgery at the lower limbs within the last twelve months prior to the measurements (d) received new AFOs in the last month.

In addition, we used two control datasets of typically developing (TD) children. These TD-databases were previously established at the clinical motion analysis laboratory consisting of an overground database including the gait data from 56 TD children (age: 11y 1mo ± 3y 10mo) and a treadmill database including the gait from 18 TD children (age: 9y 6mo ± 3y 4mo). The data of the TD children were used for the calculation of the gait indices and to express the gait variability as a percentage of the normal joint range of motion (ROM), as explained below.

Prior to participation, informed consent was given by all subjects and/or their legal guardians. The study was conducted in compliance with the principles of the Declaration of Helsinki, the principles of Good Clinical Practice and in conformity with all relevant legal regulations. Ethical approval for this study was granted by the Ethical Commission UZ/KU Leuven (CP: S65042; TD: S61520).

### Procedure

The recruited children with CP performed two repeated gait analysis sessions, with an interval of one to 14 days. All children were repeatedly assessed by the same clinician (to rule out inter-observer variability), in all four conditions (i.e., barefoot overground, barefoot treadmill, AFO overground, AFO treadmill). An overview of the study design is illustrated in Figure 1. The order of the different sessions was alternated between patients, based on availability of the overground/treadmill gait lab. In most cases (n=7) following order of conditions was maintained (a) barefoot overground (b) AFO overground (c) AFO treadmill (d) barefoot overground. Children with GMFCS-level III walked with their posture control walker in the overground setting, while they used the handrails as support during the treadmill condition.

**Figure 1.**
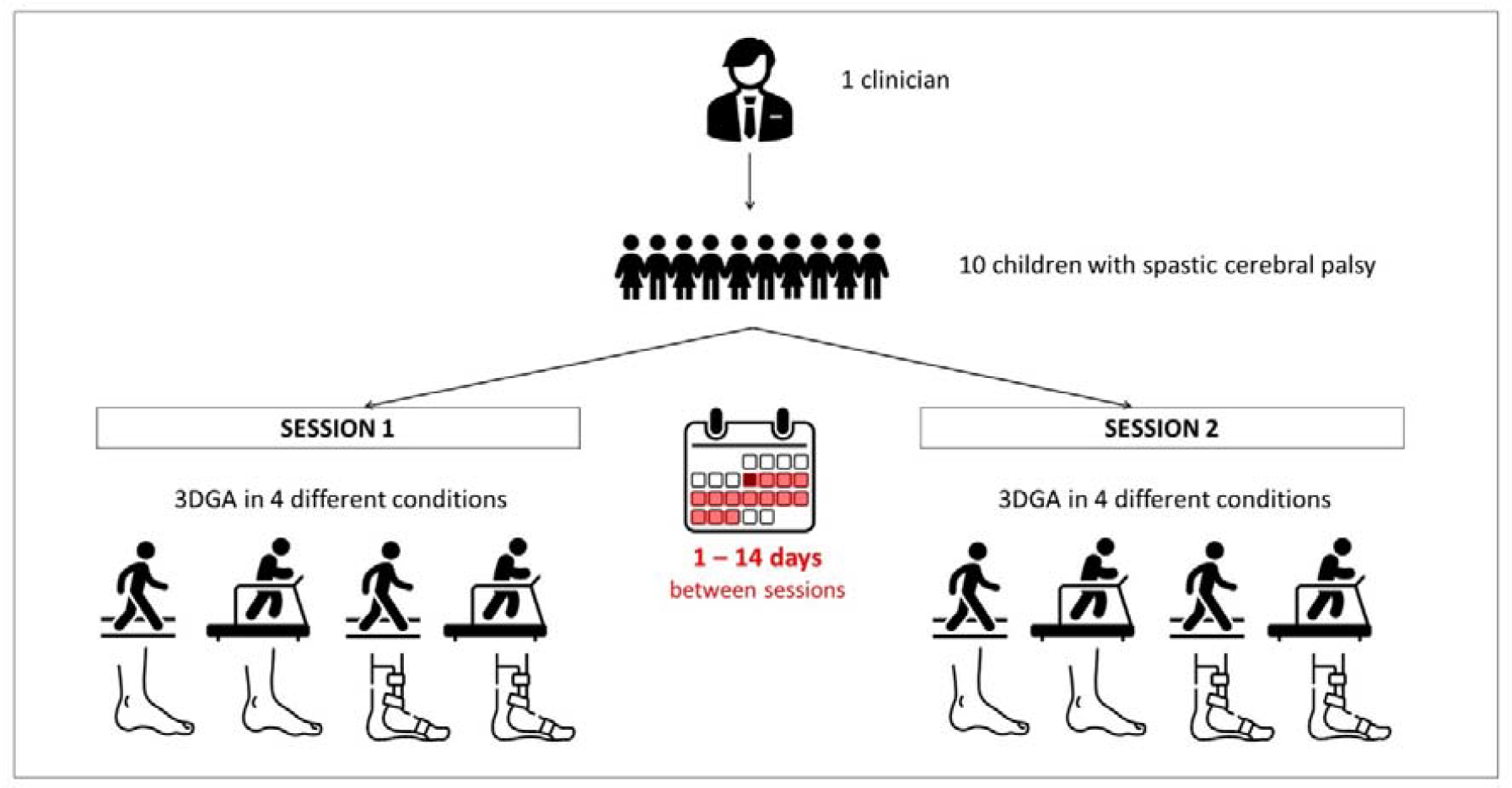
Overview of the study design. Each participant was tested by the same clinician on two separate occasions (session 1 and 2) in four different conditions: barefoot walking overground, barefoot walking on the treadmill, walking with ankle-foot orthoses overground and walking with ankle-foot orthoses on the treadmill. The interval between both sessions was one to fourteen days. Abbreviation: 3DGA = three-dimensional gait analysis

During the 3DGA, the children were asked to walk at their preferred speed during the overground conditions. For the treadmill condition, a fixed walking speed had to be chosen. This pace was initially 10% lower than during overground trials and could then be further adjusted during a barefoot habituation time. When the patient indicated a comfortable speed, the treadmill 3DGA was performed while keeping the final walking speed constant.

The 3DGAs were performed with 10 optoelectronic cameras (Vicon Motion Systems, Oxford, UK) in combination with two force plates (Advanced Mechanical Technology Inc., USA), which were integrated in a 10m walkway overground, in the overground gait laboratory and with an instrumented treadmill (GRAIL – Motek Medical, the Netherlands), in the treadmill gait laboratory. Sixteen light reflecting markers were fixed to the skin of the children according to the lower body PlugInGait marker model (Oxford metrics, Oxford, UK). At least three trials per walking condition were collected.

Using the Vicon Nexus Software (Oxford Metrics, Oxford, UK), the continuous gait data were segmented into different gait cycles, and time-normalized to the duration of the gait cycle. The kinematic and kinetic waveforms were obtained by yielding 101 points of data per curve. The kinematic data included the cardan angles (expressed in degrees) of the ankle, knee and hip and the orientation of the pelvis (pelvis data were not used in the current study), decomposed in different anatomical planes (only the sagittal plane data were used in the current study), based on the lower body PlugInGait kinematic model. The kinetic data included internal joint moments and joint power, normalized to body weight (expressed in Nm/kg and W/kg, respectively), defined through inverse dynamics, based on the kinematic and force plate data.

### Data Analysis

The analyses were performed on two types of data, namely the discrete gait indices as well as the continuous gait waveforms. The gait indices GPS and GVS of the three lower-limb joints were calculated^3^ . Kinematic and kinetic continuous waveforms of 101 datapoints were extracted for each participant. Using a custom Matlab® script, the quality of the data was visually checked by observing the overlaying trials, and cycles with outliers or obvious marker placement issues (i.e., 50% of the gait cycle exceeding the 2 standard deviations from the average gait cycle) were excluded. The first ‘good’ 10 gait cycles of kinematic and kinetic data were selected for further analysis for each participant, except for the overground walking condition, where only 3 kinetic gait cycles per child were available. Kinetic data was not collected from participants who utilized a posture control walker (GMFCS-level III, N=3) during the overground condition. Only the participant’s most involved side, based on the highest Modified Ashworth Scale score of m. gastrocnemius, was used for further analysis.

### Statistical Analysis

Separate analyses were performed on the two types of data. First, for the statistical analysis of the gait indices (i.e., GPS and GVS), the inter-trial and -session variability was defined by the use of the intra-class correlation coefficient (ICC; 2,1) (two-way random model with absolute agreement), along with the 95% confidence interval and the standard error of measurement (SEM). The SEM is the standard deviation of several measurements made on the same participant.

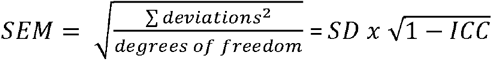 where SD is the standard deviation of the grand mean (mean of session 1 and session 2) from all participants.

Furthermore, the SEM percentage was computed in respect to the total ROM of the lower limb joints of the TD children.

The primary advantage of the SEM lies in providing a direct indication of measurement error in the same units as the original measurement, which makes the use of SEM particularly clinically relevant. Secondly, was the SEM also used as a statistical analysis for determining the inter-trial and -session variability of the kinematic and kinetic continuous waveforms in the sagittal plane. Thereby, the SEM was calculated timepoint-by-timepoint from the continues waveforms, according to the methods of Schwartz et al.^5^ . Figure 2 represents a graphical view of the inter-trial and inter-session SEM for a single subject. Both the continuous waveforms and the mean values of these waveforms will be reported.

**Figure 2.**
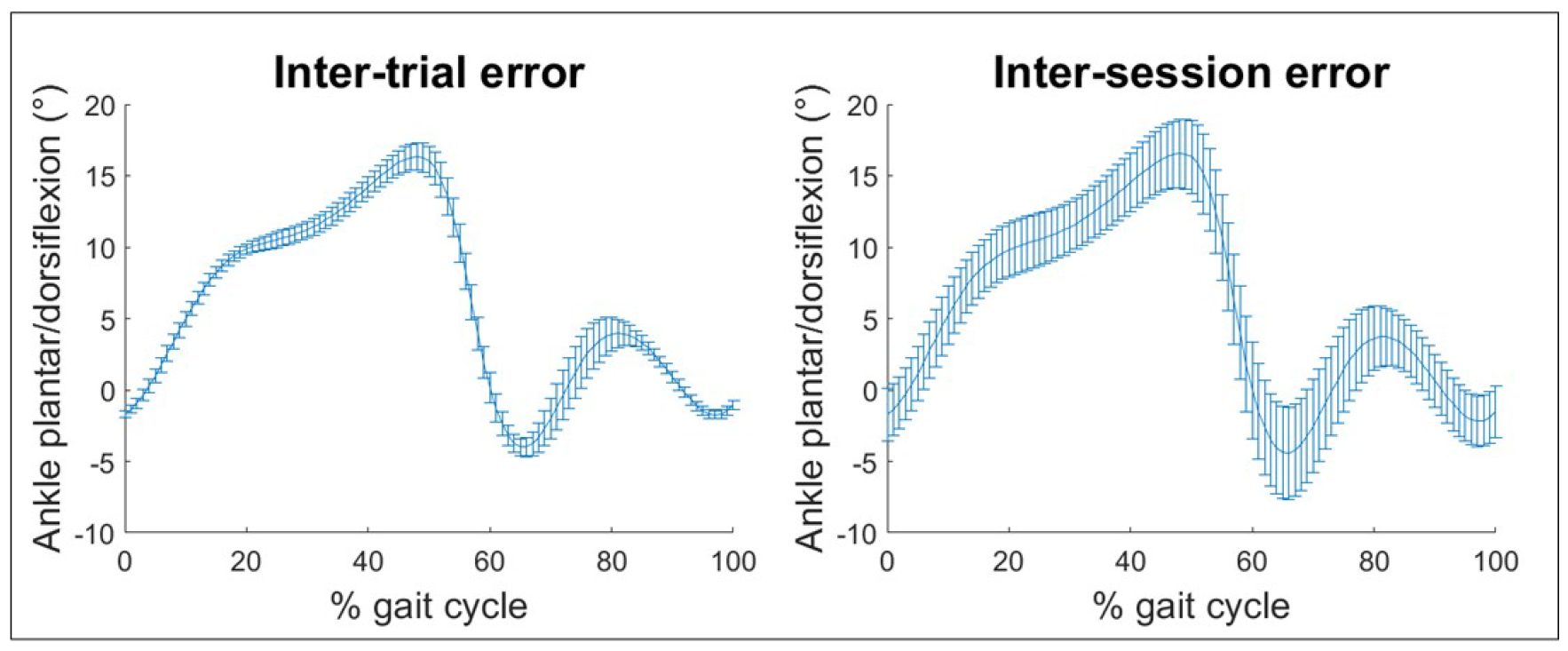
Schematic representation of the statistical analysis of the sagittal kinematic ankle data of one representative participant. Inter-trial differences (left graph) arise due to the natural (intrinsic) variability that occurs from stride-to-stride. Inter-session errors (right graph) indicate how well a single clinician consistently gathers gait data.

## Results

For all conditions, between good to excellent ICC-values were found for the inter-session GPS (ICC = 0.85 - 0.98) and GVS (0.85 - 0.99) of the different joints (Table 1). The inter-trial ICC-values were all found to be excellent (ICC = 0.99) for the GPS and GVS. The GVS of all three joints showed favourable SEM-values in all conditions, with the knee joint displaying the highest inter-trial errors (SEM = 0.31° - 0.58°), while the hip joint showed the highest inter-session errors (SEM = 0.52° – 1.11°). Overall, a higher SEM for the inter-session measurements compared to the inter-trial measurements was found. When considering the SEM as a percentage of the total TD ROM, the three lower limb joints exhibited comparable inter-trial values (%SEM=0.5%-1.3%). However, the knee displayed the lowest inter-session values (%SEM=0.5%-1.8%), whereas the ankle joint demonstrated the highest inter-session values (%SEM=0.9%- 3.0%).

**Table 1.** Variability values for the Gait Profile Score and the Gait Variable Scores of the hip, knee and ankle

| Measure | Condition | Inter-trial |  | Inter-session |  |
| --- | --- | --- | --- | --- | --- |
|  |  | ICC (95% CI) | SEM (%ROM) | ICC (95% CI) | SEM (%ROM) |
| **GPS** | AFO overground | 0.99 (0.98-0.99) | 0.32° | 0.98 (0.89-1.00) | 0.45° |
|  | AFO treadmill | 0.99 (0.98-0.99) | 0.25° | 0.85 (0.05-0.98) | 0.91° |
|  | BF overground | 0.99 (0.99-1.00) | 0.31° | 0.95 (0.80-0.99) | 0.68° |
|  | BF treadmill | 0.99 (0.99-1.00) | 0.19° | 0.88 (0.27-0.98) | 0.61° |
| **GVS**<br>hip flexion | AFO overground | 0.99 (0.99-1.00) | 0.51° (1.1%) | 0.95 (0.73-0.99) | 1.11° (2.4%) |
|  | AFO treadmill | 0.99 (0.98-1.00) | 0.32° (0.8%) | 0.97 (0.80-0.99) | 0.54° (1.4%) |
|  | BF overground | 0.99 (0.99-1.00) | 0.46° (1.0%) | 0.94 (0.75-0.99) | 1.03° (2.2%) |
|  | BF treadmill | 0.99 (0.98-1.00) | 0.27° (0.7%) | 0.96 (0.80-0.99) | 0.52° (1.4%) |
| **GVS**<br>knee flexion | AFO overground | 0.99 (0.99-1.00) | 0.42° (0.7%) | 0.98 (0.92-1.00) | 0.59° (1.0%) |
|  | AFO treadmill | 0.99 (0.99-1.00) | 0.58° (1.0%) | 0.97 (0.82-1.00) | 1.01° (1.8%) |
|  | BF overground | 0.99 (0.99-1.00) | 0.31° (0.5%) | 0.99 (0.97-1.00) | 0.30° (0.5%) |
|  | BF treadmill | 0.99 (0.98-1.00) | 0.48° (0.8%) | 0.98 (0.91-1.00) | 0.68° (1.2%) |
| **GVS**<br>ankle dorsiflexion | AFO overground | 0.99 (0.99-0.99) | 0.25° (0.8%) | 0.85 (0.27-0.97) | 0.90° (3.0%) |
|  | AFO treadmill | 0.99 (0.98-1.00) | 0.22° (0.9%) | 0.89 (0.02-0.99) | 0.62° (2.5%) |
|  | BF overground | 0.99 (0.98-1.00) | 0.38° (1.3%) | 0.95 (0.73-0.99) | 0.82° (2.7%) |
|  | BF treadmill | 0.99 (0.99-1.00) | 0.23° (0.9%) | 0.99 (0.97-1.00) | 0.22° (0.9%) |

*Abbreviations: ICC = Intraclass correlation coefficients; CI = confidence interval; SEM = standard error of measurement; ROM = range of motion; GPS = Gait Profile Score; GVS = Gait Variable Score; AFO = ankle-foot orthosis; BF = barefoot*

For the gait waveforms, the SEM for each timepoint of the gait cycle of the inter-trial and -session kinematics and kinetics are presented in Figure 3. Overall, no differences in SEM were observed between conditions for kinematics, except for the ankle joint, where we observed different SEM-values between the AFO and barefoot condition. Here, the AFO resolved in smaller SEM-values (mean inter-trial AFO = 0.14°; mean inter-trial BF = 0.55°; mean inter-session AFO = 1.12°; mean inter-session BF = 2.22°), most notable during the swing phase. The ankle joint showed the lowest SEM-values (mean inter-trial = 0.35°; mean inter-session = 1.67°), and the knee joint displayed the highest SEM (mean inter-trial = 0.61°; mean inter-session = 3.53°). For all the kinetic parameters, the treadmill condition demonstrated smaller SEM-values in comparison to the overground condition. The hip revealed the highest SEM-values for joint moments (mean inter-trial hip = 0.041Nm/kg; mean inter-trial knee = 0.026Nm/kg; mean inter-trial ankle = 0.036Nm/kg; mean inter-session hip = 0.143Nm/kg; mean inter-trial knee = 0.097Nm/kg; mean inter-trial ankle = 0.119Nm/kg) and power (mean inter-trial hip = 0.065W/kg; mean inter-trial knee = 0.051W/kg; mean inter-trial ankle = 0.037W/kg; mean inter-session hip = 0.206W/kg; mean inter-trial knee = 0.181W/kg; mean inter-trial ankle = 0.186W/kg).

**Figure 3.**
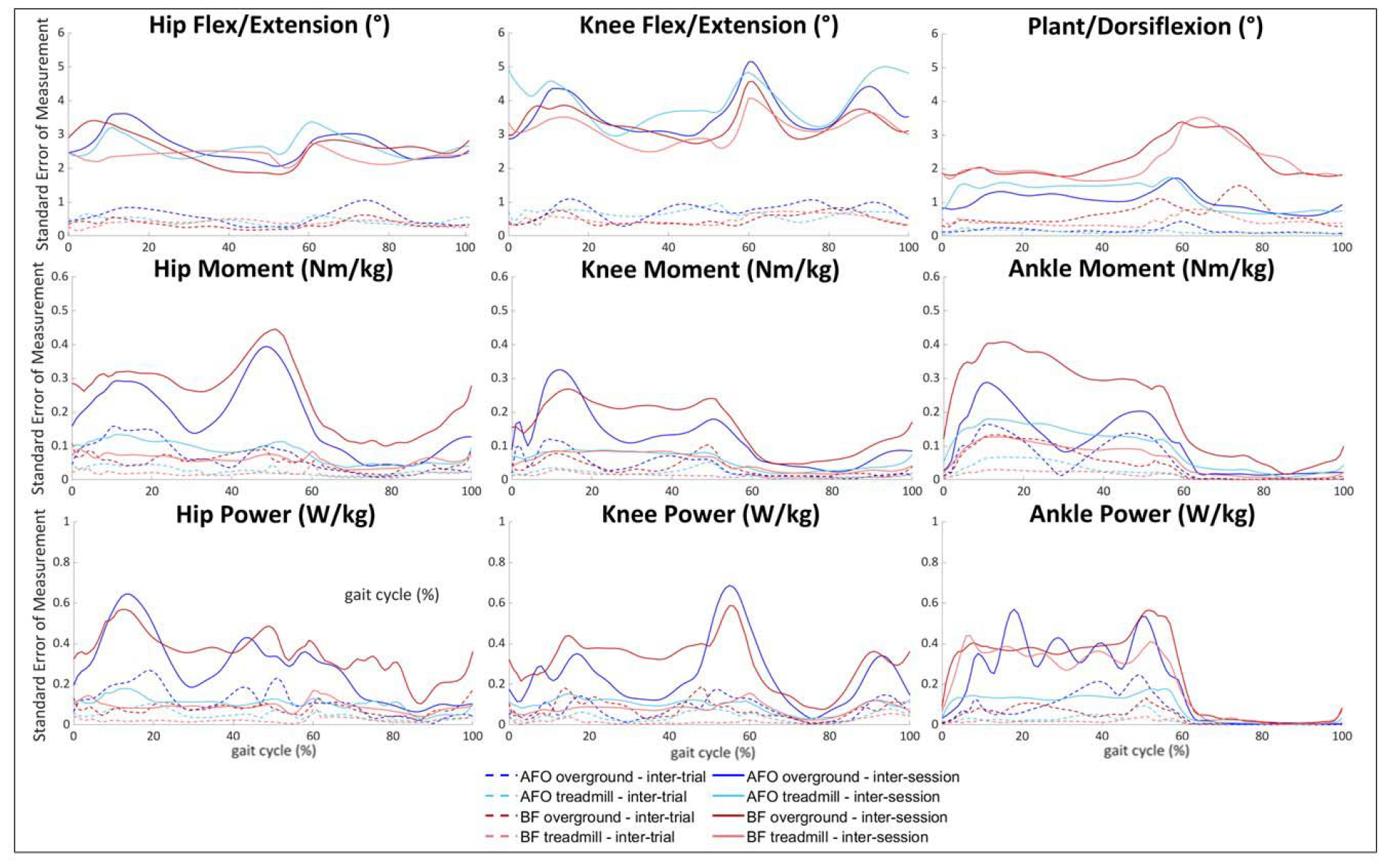
Standard error of measurement of the kinematics (1^st^ row) and the kinetics (moments – 2^nd^ row; powers – 3^rd^ row) for each point of the gait cycle for the hip (1^st^ column), the knee (2^nd^ column) and the ankle (3^rd^ column) defined for children with spastic cerebral palsy (N=10).

## Discussion

Overall, good to excellent repeatability of 3DGA was found for children with spastic CP who walked in four conditions (i.e., overground, on the treadmill, barefoot and with AFOs).

We first quantified the repeatability of the barefoot overground condition, as this is traditionally the most used condition in gait analysis. For the GPS and GVS, excellent inter-trial ICC values were obtained (GPS: ICC = 0.95 and GVS: ICC = 0.94 – 0.99) in all measured joints (sagittal plane). Rasmussen et al.^12^ found slightly better (higher) ICC values, but worse (higher) SEM-values for both the GPS and the GVS. This might be explained by the slightly more involved group of participants (3 children with GMFCS-level III) in the current study, who tend to have higher scores on the GPS and GVS in comparison to children with GMFCS-level I to II. For the inter-trial measurements, our results for the hip (mean SEM = 0.34°), knee (mean SEM = 0.50 °) and ankle (mean SEM = 0.67°) were slightly smaller or almost similar to those reported by Schwartz et al.,^5^ which were respectively 1.17°, 0.60° and 0.65°. For inter-session repeatability, the SEM-values of the current study were in line with the results reported by Schwartz et al.^5^ It should be noted that the latter study was performed on a different population (i.e., two healthy adults). In the current study, the knee emerged as the most consistent joint, demonstrating the lowest SEM-values expressed as percentage of the TD total range of motion (ROM). Contextualizing these findings with respect to the joint’s overall ROM is crucial. Without such contextualization, the ankle joint might erroneously appear to possess the lowest SEM-values. However, it is important to note that the ankle has the lowest ROM of all three joints and thus the smallest range for errors.

As a second step, we compared the 3DGA repeatability between the barefoot and AFO-condition. Overall, for kinematic data, rather similar SEM-values were observed between the two conditions, except for the ankle, where the AFO showed lower SEM-values (mean inter-session SEM AFO = 1.12°; mean inter-session SEM barefoot = 2.22°), most notable during the swing phase (Figure 3). This can be explained by the restriction of ankle motion introduced by the AFO. For the kinetics and the gait deviation indices, no obvious differences were found between the repeatability for the barefoot and AFO-condition.

Finally, the repeatability between the treadmill and overground condition was compared. The overground condition showed the highest ICC values (inter-session ICC = 0.95-0.98), while the treadmill condition showed lower but still good ICC values (inter-session ICC = 0.85-0.88). The joint-specific kinematic analyses highlighted that the overground-condition had larger SEM-values for the GVS of the hip and the ankle in comparison to the treadmill-condition, while the opposite was observed for the knee GVS. For the kinetic parameters, the treadmill condition had lower SEM-values than the overground condition, indicating more repeatable kinetics (Figure 3). This might be explained by the continuous walking speed on the treadmill in comparison to the self-selected walking speed overground. In addition, in the overground condition, due to practical reasons, the kinetic outcomes represented the average of only 3 trials, which was much lower than the 10 trials used for the treadmill condition.

The study results are relevant for different purposes. First, the quantified consistency between gait sessions can be used as a reference for judging patient-specific repeated gait sessions in children with CP and can thus support the interpretation of routine clinical 3DGA. Indeed, repeated 3DGA are often used in children with CP to outline their natural course of gait deviations, to plan and finetune treatments and to define the effect of treatments. Secondly, gait may become inconsistent in new walking conditions, such as walking with a new orthosis. The current results thereby provide a reference to define if a patient got well-adapted to the new walking condition. Finally, the results can also be used as a reference to evaluate newly trained gait analysis assessors.

### Study limitations

First, our current sample size did not allow any subgroup analyses (i.e., per GMFCS-level, type of orthoses, gait pattern etc.). Secondly, outliers were identified and removed from the dataset, enhancing the consistency of the selected gait data. It is important to note that our findings can only be generalized to similar datasets (i.e., not derived from a single gait cycle). Moreover, our results are gait laboratory and clinician specific. Thirdly, compared to the kinematics, the kinetic dataset during overground walking was smaller because we exclusively utilized kinetic overground data from children who were walking without walking aids (GMFCS-level I-II, N=7 children). Additionally, we limited the analysis to three gait cycles in the overground condition, because of the increased difficulty for children to correctly hit the overground force plates (which was not an issue while walking on the treadmill with embedded force plates). Finally, the SEM-values were put in perspective to the total ROM of the joint of TD children from our established overground and treadmill database. Because this database only included the barefoot condition, the AFO SEM-values were also compared to the joint ROMs in barefoot condition.

### Conclusions

This study showed that performing a 3DGA on an overground walkway while walking barefoot results in reliable gait parameters in children with CP. Moreover, also other walking conditions showed good to excellent 3DGA repeatability. For kinematics, almost no differences were found between the conditions, except for the ankle which showed smaller SEM-values when the participants were walking with their AFOs in comparison to barefoot walking. Taking the total joint ROM into account, the knee joint was the most repeatable joint, while the ankle had the highest %SEM-values. For kinetics, treadmill conditions showed better repeatability than the overground conditions.

## Data Availability

All data produced in the present study are available upon reasonable request to the authors

